# Evaluating Large Language Models for Automatic Detection of In-Hospital Cardiac Arrest: Multi-Site Analysis of Clinical Notes

**DOI:** 10.1101/2025.08.04.25331524

**Authors:** Uğurcan Vurgun, Aarthi Kaviyarasu, Sy Hwang, Ashley Batugo, Sunil Thomas, Brandon Tang, Ana Acevedo, Oscar Mitchell, Danielle L. Mowery

**Affiliations:** Department of Biostatistics, Epidemiology, & Informatics; Department of Emergency Medicine, Division of Pulmonary, Allergy, and Critical Care, Center for Resuscitation Science University of Pennsylvania Philadephia, PA, 19104, United States

**Keywords:** In-Hospital Cardiac Arrest, Large Language Models, Healthcare AI

## Abstract

In-hospital cardiac arrest (IHCA) affects over 200,000 patients annually in the United States, yet its detection through manual chart review remains resource-intensive and often delayed. We evaluated the performance of four open-source large language models (LLMs) and GPT-4o in identifying IHCA cases from 2,674 clinical notes across five hospitals. While GPT-4o achieved the highest performance (F1-score: 0.90, recall: 0.97), several open-source models demonstrated comparable capabilities, suggesting their viability for clinical applications. Our systematic analysis of model outputs revealed that performance was strongly influenced by site-specific documentation practices, with inter-site agreement rates varying by over 20%. Through detailed error analysis, we identified key challenges including medical terminology hallucinations and structural inconsistencies in model reasoning. These findings establish a framework for implementing LLM-based IHCA detection systems while highlighting critical considerations for their clinical deployment.

## 1. Introduction

Accurate and timely reporting of IHCA events is crucial to facilitate quality improvement (QI) initiatives, such as reviewing cardiopulmonary resuscitation (CPR) quality, optimizing team performance, and benchmarking IHCA outcomes. Although care teams can readily recognize IHCA during the event itself, subsequent reporting of these events after the event often lacks consistency and poses a major barrier to QI and research efforts. Reliance on administrative billing codes or human-driven processes lacks precision, can be costly, and can delay or miss critical data collection.^1,2^ This issue is particularly acute during the night and weekends, when nursing managers and other key champions may not be available, and during periods of staff shortages and turnover. LLMs provide capabilities for interpreting unstructured clinical text, reducing human variability, and streamlining data extraction.^3^ Yet, their validity in critical care depends on rigorous testing of accuracy, robustness, and instruction adherence.

## 2. Related Work

Traditional IHCA identification methods rely on billing codes, such as the International Classification of Diseases (ICD), or manually-created QI and research registries, which often lack timeliness and accuracy.^1,2^ Previous works found that using ICD-9 codes to identify cardiac arrest patients lacked both sensitivity and specificity, with one large registry study reporting that over 36% of IHCA events were not identified through billing and procedure code analyses.^1,2^ Consistent with these works, our group found the use of ICD-10 codes to have low sensitivity for the identification of IHCA events from a QI database, with 32% of events being missed,^4^ demonstrating a need for novel approaches to accurately identify IHCA and facilitate research and QI efforts.

Recent advancements highlight the potential of AI in medical text analysis, including the use of LLMs to interpret complex notes.^5,6^ Luo et al.^7^ introduced BioGPT, a generative model pre-trained on biomedical corpora, and had shown promise in extracting nuanced clinical information and summarizing patient records. Singhal and colleagues^8^ demonstrated that foundational LLMs could effectively disambiguate medical terminology without the need for domain-specific fine-tuning. Rapid advancements have enabled practical applications across various medical domains, including identifying within clinical narratives cognitive decline in the elderly,^9^ headache frequency in neurology consultations,^10^ and sleep patterns in Alzheimer’s disease patients.^11^ In this pilot study, we conducted a summative evaluation to assess how well LLMs could discern positive IHCA cases from negative IHCA cases and a formative evaluation to understand conformance with IHCA criterion.

## 3. Methodology

### 3.1. Data Collection and Preprocessing

We collected 2,674 clinical notes from 684 patients from 5 hospitals from southeastern Pennsylvania, emphasizing discharge summaries, progress notes, plan-of-care notes, and assessments. To protect patient privacy, notes were de-identified using PHIlter.^12^ Domain experts annotated each note- and patient-level labels to identify features relevant to IHCA, including patient demographics, comorbidities, vital signs, and events preceding the arrest. Double annotation with adjudication by a senior clinician ensured labeling consistency. Patient-level labels were assigned as IHCA-positive when there was documented evidence of a cardiac arrest occurring during an inpatient hospital stay, requiring chest compressions or advanced cardiac life support interventions. Cases were labeled as IHCA-negative when there was no documented cardiac arrest or when cardiac arrests occurred in outpatient settings such as clinics or emergency departments prior to admission. For model evaluation, we provided structured instructions requiring verification of three hierarchical criteria: (1) explicit documentation of cardiac arrest, (2) confirmation that the event occurred in an inpatient setting after admission, and (3) documentation of resuscitative interventions including chest compressions or advanced cardiovascular life support (ACLS) protocols. Note-level labels were derived from patient-level annotations, with notes documenting the IHCA event or its immediate aftermath labeled as positive, enabling temporal analysis of documentation patterns.

### 3.2. Model Selection and Evaluation

We evaluated five LLMs: GPT-4o and four open-source alternatives (Mistral-Nemo-Instruct-2407, Meta-Llama-3.1-8B-Instruct, princeton-nlp/gemma-2-9b-it-SimPO, and THUDM/glm-4-9b-chat). All models were applied with their base configuration without additional fine-tuning, employing a zero-shot learning approach with chain-of-thought prompting. Models were evaluated using an identical prompt and generation parameters (temperature=0.2) to ensure fair comparison:

Given the patient’s notes summary, assess whether the patient had an in-hospital cardiac arrest (IHCA) event. IHCA refers to a cardiac arrest that occurs in an inpatient setting where the patient is admitted and receiving care. Cardiac arrests occurring in outpatient settings, such as the hospital’s cafeteria or during a visit, do not count as IHCA. Cardiac arrest refers to the sudden loss of cardiac function leading to a lack of a pulse or signs requiring that the patient receives chest compressions, or ACLS.

Steps to determine IHCA:

1. Identify if there is a mention of cardiac arrest.
2. Determine if the cardiac arrest occurred in an inpatient setting.
3. Confirm if the patient received chest compressions.

Respond with ‘positive’ or ‘negative’ and give your rationale.

### 3.3. Summative Evaluation Metrics

To determine how well LLMs discerned IHCA positive from IHCA negative cases, we evaluated model performance using standard binary classification metrics at the patient-level: F1-score, balanced accuracy, precision, and recall. To assess statistical robustness, we computed 95% confidence intervals using non-parametric bootstrapping with 100,000 resamples. Additionally, we measured instruction prompt following performance through a binary assessment of each model response: responses were considered compliant if they (1) began with an explicit ‘positive’ or ‘negative’ classification and (2) provided a rationale referencing the required IHCA criteria. We aggregated these assessments into an instruction following score, calculated as the proportion of responses meeting both criteria across all cases.

### 3.4. Formative Evaluation Metrics

To better understand the rationale of IHCA classification and conformance with IHCA criterion, we randomly sampled and manual reviewed up to 5 patients and their subsequent 4 notes from each of the concordant (true positives and negatives) and dis-concordant (false positives and negatives) cells of the contingency matrix from each of the 5 hospitals. Three reviewers with clinical expertise (AK, BT, AA) evaluated the reliability of the responses from the LLM at a note-level using criterion of **coverage, coherence, relevance**, and **hallucinations** defined in Table 1. Coverage and coherence were measured by 5-point Likert scales; relevance were measured by a 3-point scale. Review and evaluation was conducted through REDCap, an encrypted, HIPAA-compliant, and hospital-approved data management software.^13^

**Table 1.** Evaluation criteria and scales used by annotators.

| Criteria | Question | Evaluation Scale |
| --- | --- | --- |
| Coverage | Does the rationale accurately represent the information in the original text? | • **5** – Yes; the rationale covers all the important information present in the original text.<br>• **4** – Mostly; the rationale covers most, but not all of the important information.<br>• **3** – Somewhat; the rationale covers some of the important information, but also misses some of them.<br>• **2** – Not really; the rationale misses most of the important information.<br>• **1** – No; the rationale does not cover any of the important information present in the original text. |
| Coherence | Is the rationale coherent? | • **5** – Yes; the rationale is easy to read and understand.<br>• **4** – Mostly; the rationale is readable, but not straightforward to understand.<br>• **3** – Somewhat; the rationale is readable, but confusing.<br>• **2** – Not really; the rationale has some grammatical errors or non sequiturs.<br>• **1** – No; the rationale is unintelligible or incomprehensible. |
| Relevance | Does the rationale answer the original question? | • **3** – Yes; the rationale answers the original question.<br>• **2** – Partially; the rationale answers the original question, but not fully.<br>• **1** – No; the rationale does not answer the original question. |
| Hallucination | Does the rationale contain information not present in the original text? | • **0** – No; the rationale does not contain information not present in the original text.<br>• **1** – Yes; the rationale contains information not present in the original text. |

## 4. Results

We evaluated five LLMs on 2,674 clinical notes from 684 unique patients. Each patient contributed up to four notes, primarily discharge summaries, progress notes, plan-of-care notes, and assessments, chosen for their frequent documentation of IHCA events.

### 4.1. Summative Evaluation: LLM Classification at Patient-level

Figure 1 summarizes each model’s performance metrics. GPT-4o obtained the highest F1-score (0.90 *±* 0.01), along with a balanced accuracy of 0.90 *±* 0.01 and a 0.97 recall for IHCA cases. Several open-source LLMs exhibited competitive performance: Mistral-Nemo-Instruct-2407 (F1: 0.84 *±* 0.05), princeton-nlp/gemma-2-9b-it-SimPO (F1: 0.82 *±* 0.07), THUDM/glm-4-9b-chat (F1: 0.79 *±* 0.00), and Meta-Llama-3.1-8B-Instruct (F1: 0.79 *±* 0.05). Confidence intervals were derived through non-parametric bootstrapping with 100,000 resamples to assess robustness.

**Fig. 1.**
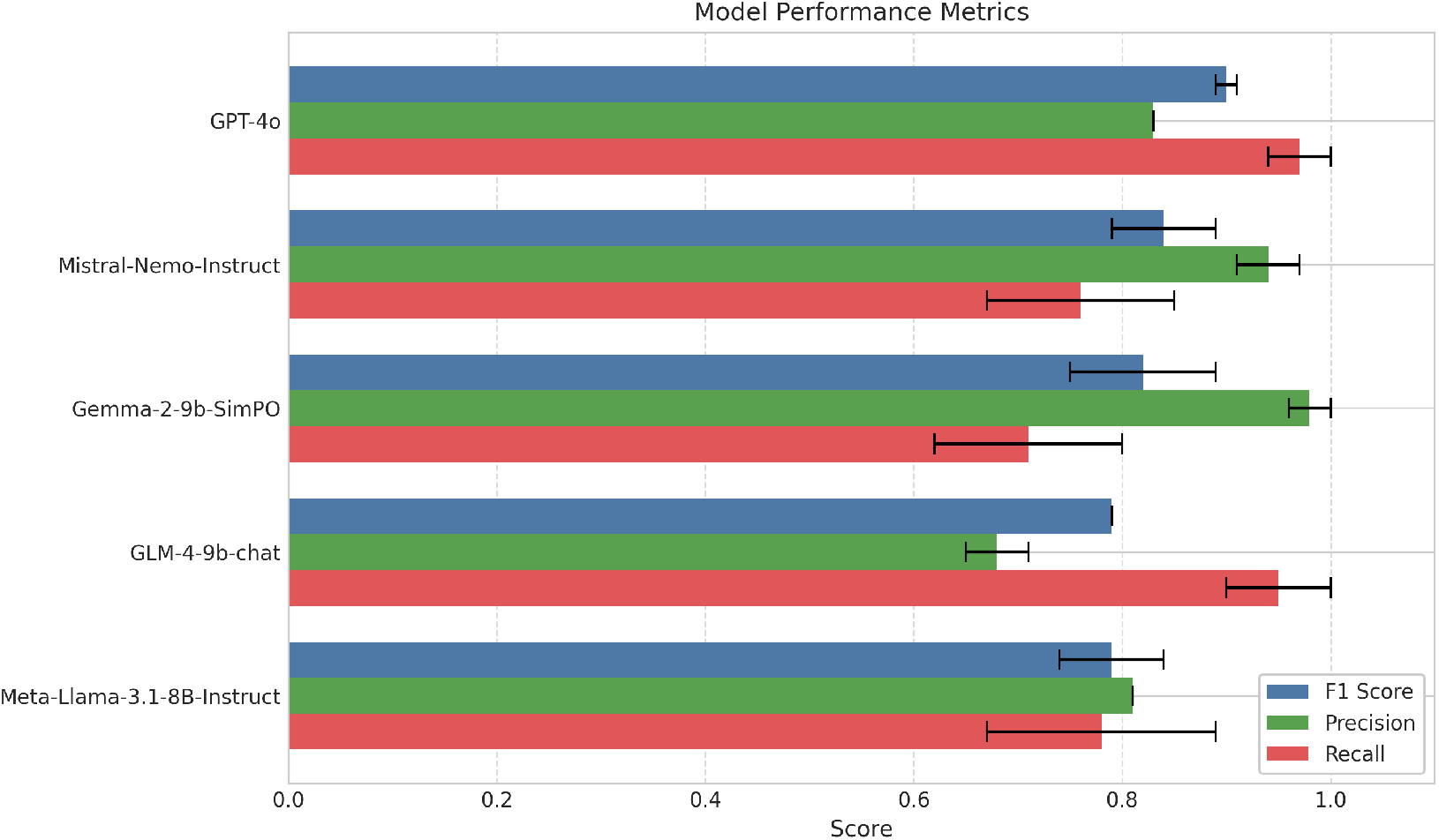
Performance comparison of five LLMs on IHCA detection, including F1-score, precision, and recall with confidence intervals.

### 4.2. Formative Evaluation: Classification and Rationale Subanalysis on Note-level

To evaluate not just binary predictions but also rationale quality, focusing on coverage, coherence, relevance, and hallucination detection, we selected a subset of 256 clinical notes from 64 patients across five hospitals from the Llama-3.1-8B-Instructmodel for annotation. We selected this model because of its high F1-score and high instruction following capabilities which leads itself nicely to a human-in-the-loop review model. Each clinical note was evaluated independent of any classifications at the patient-level, as well as the other three notes from the same patient record. Table 1 shows the criteria and the evaluation scales adapted from Sudeshna et al.’s study.^14^

The analyzed notes predominantly comprised of progress notes (67.2%, n=172) and discharge summaries (26.6%, n=68). Figure 2 illustrates the distribution of IHCA indicators in these clinical notes. Explicit cardiac arrest mentions were most frequent, followed by inpatient setting documentation. Chest compression documentation appeared less commonly, while a small fraction of notes contained no clear IHCA indicators.

**Fig. 2.**
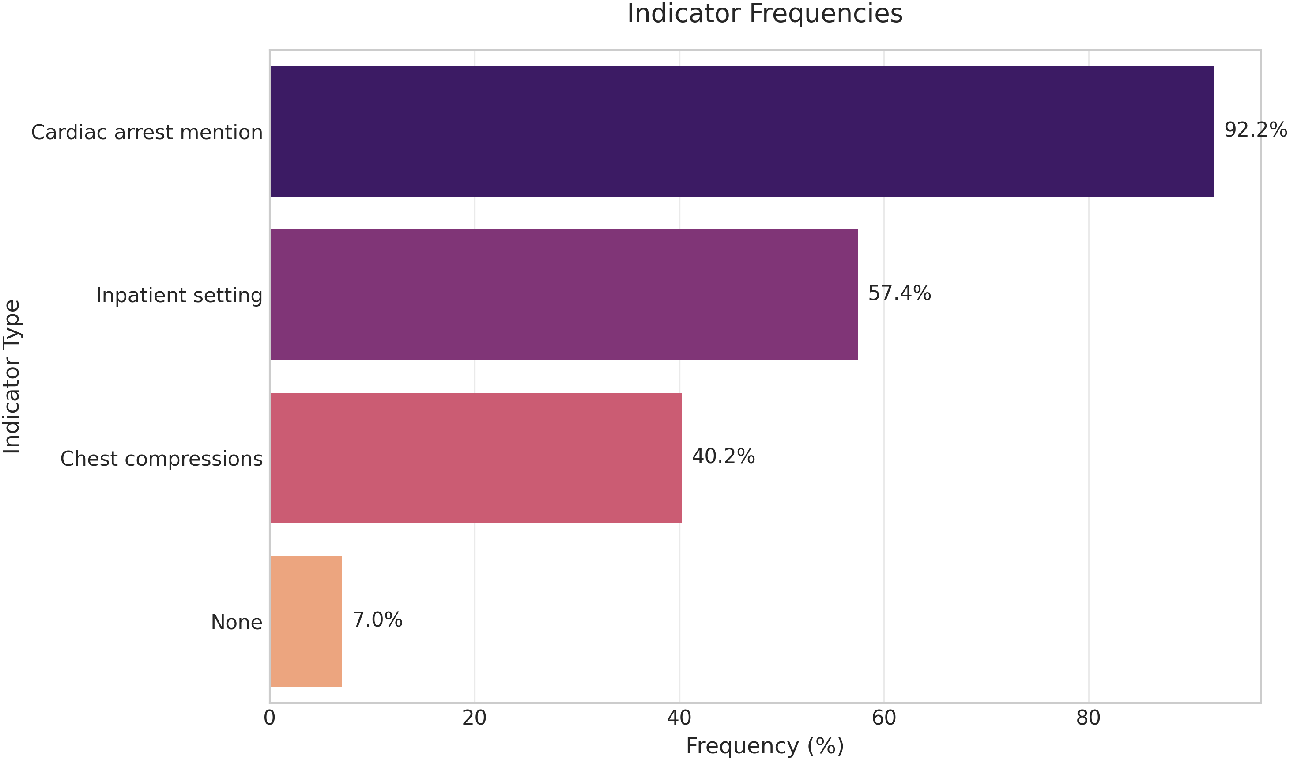
Distribution of IHCA indicators in clinical notes from the annotated dataset.

Figure 3 shows the agreement rates between human observations and model inferences for key IHCA indicators. The capacity of LLMs to accurately interpret IHCA indicators varied, with high detection rates for explicit cardiac arrest mentions (*>* 90%), but lower agreement for inpatient status (55%) and chest compressions (38%). Overall agreement rates also varied substantially across hospital sites: Sites 2 and 3 achieved 71.4% agreement, Site 4 70.0%, Site 5 53.8%, and Site 1 only 50.0% (not shown).

**Fig. 3.**
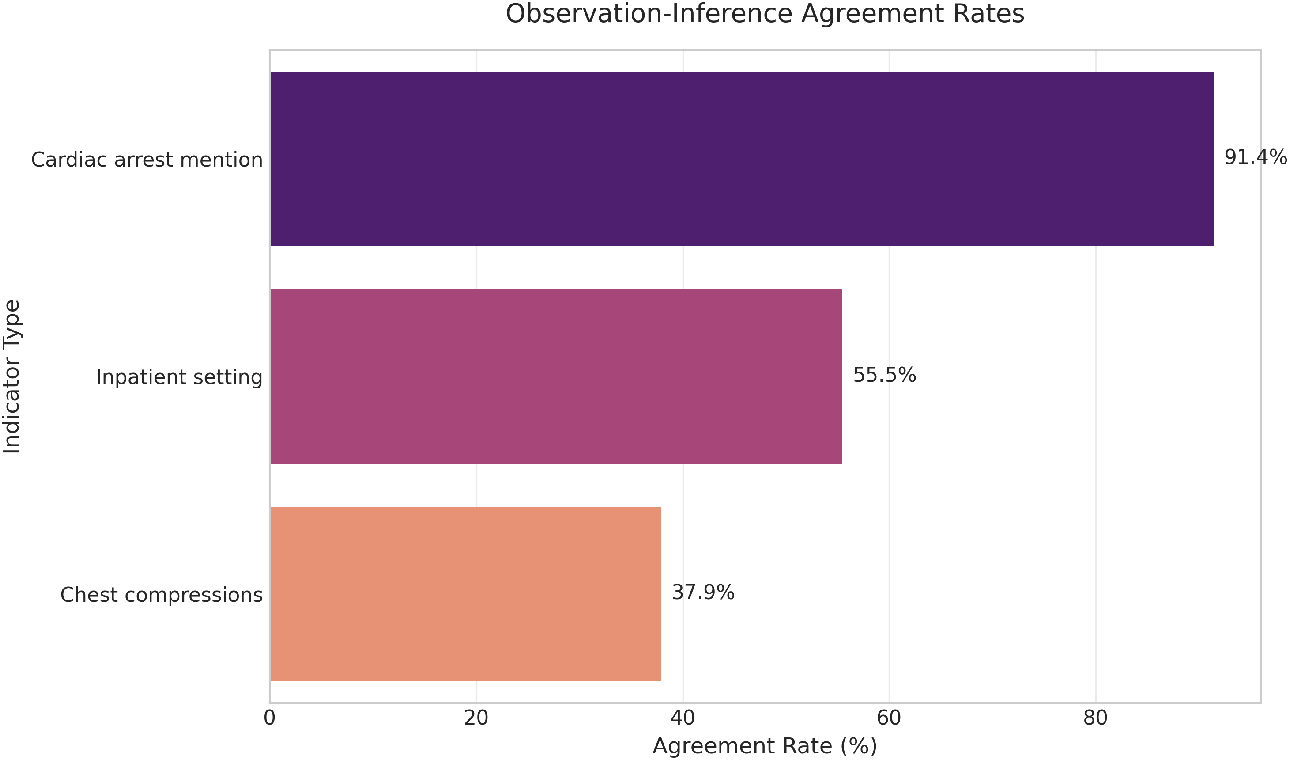
Agreement rates between human observations and model inferences for key IHCA indicators.

Figure 4 displays the distribution of coverage and coherence ratings across reviewed model outputs. Here, coverage indicates the model’s ability to capture critical IHCA-related information, and coherence reflects the readability and structural quality of the outputs. Most cases scored 5 indicating high coverage of criteria (83.6%) and readability (85.9%).

**Fig. 4.**
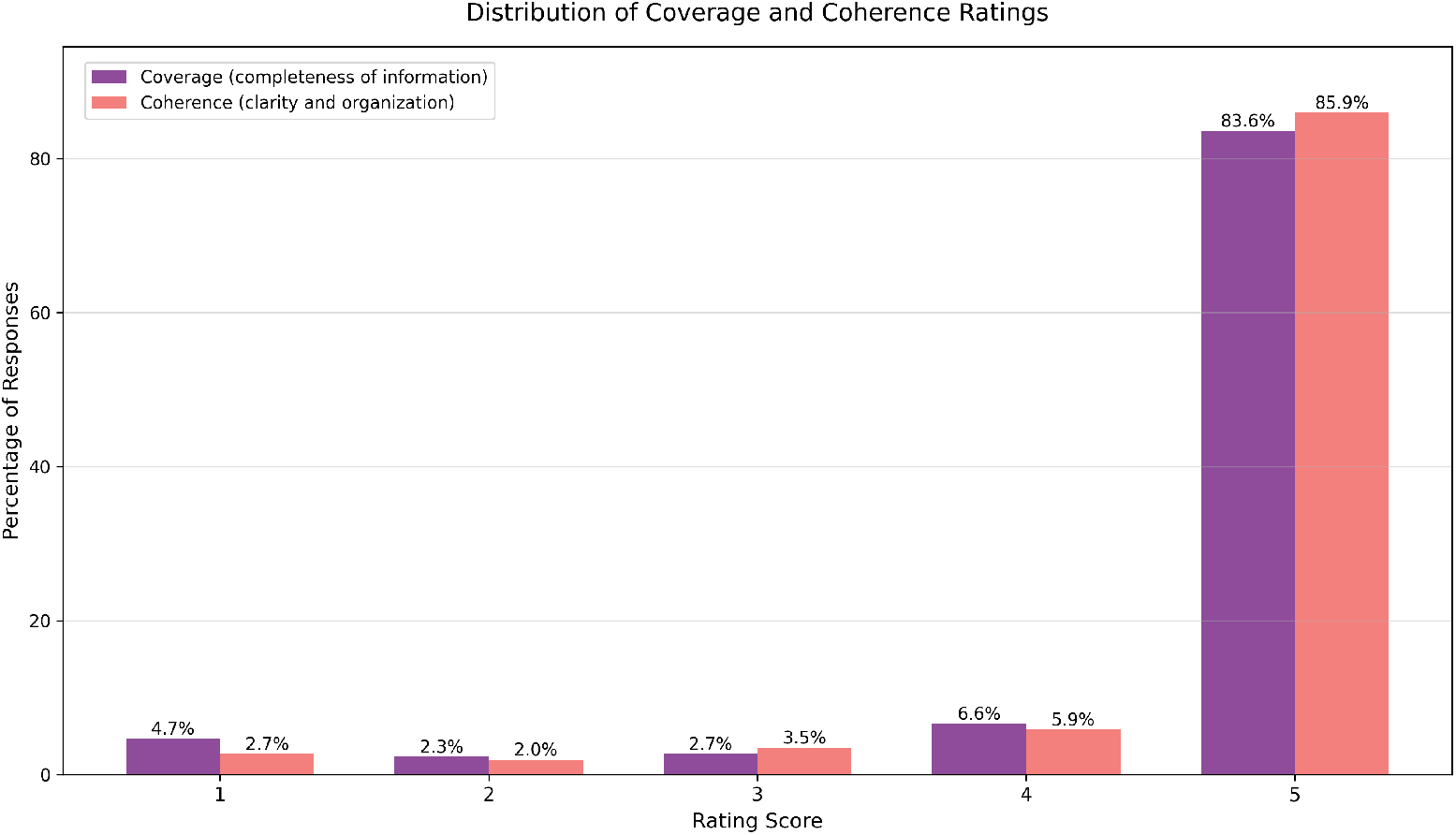
Distribution of coverage and coherence ratings across reviewed model outputs.

Figure 5 presents the hallucination rates across the three most common clinical note types. Hallucination rates varied by note type, with plan of care showing the highest rate (37.5%), discharge summaries the second highest (19.1%), and progress notes the lowest (12.8%).

**Fig. 5.**
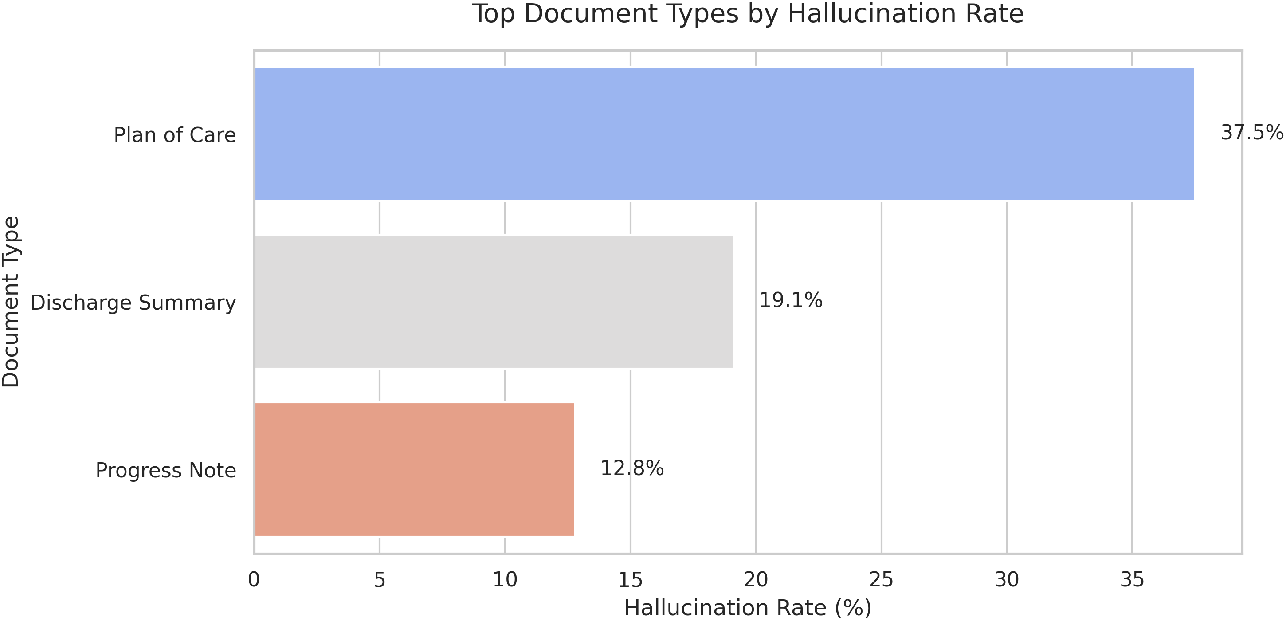
Analysis of hallucination rates across the three most common clinical note types.

## 5. Discussion

### 5.1. Summative Evaluation: LLM Classification at Patient-level

When selecting an LLM for operations, its important to not only consider classification accuracy, but also costs associated with deployment i.e., GPT-4o costs are calculated by tokens which can be computationally expensive compared to open-weight models. Although GPT-4o produced the highest performance, open-weight LLMs could produce comparable or even better recall or precision.

### 5.2. Formative Evaluation: Classification and Rationale Subanalysis on Note-level

Rationales are critical to explainability and trust in LLM predictions. The feedback notes written by our expert annotators further reveal several error types in the model’s outputs. These errors are summarized in Table 2.

**Table 2.**
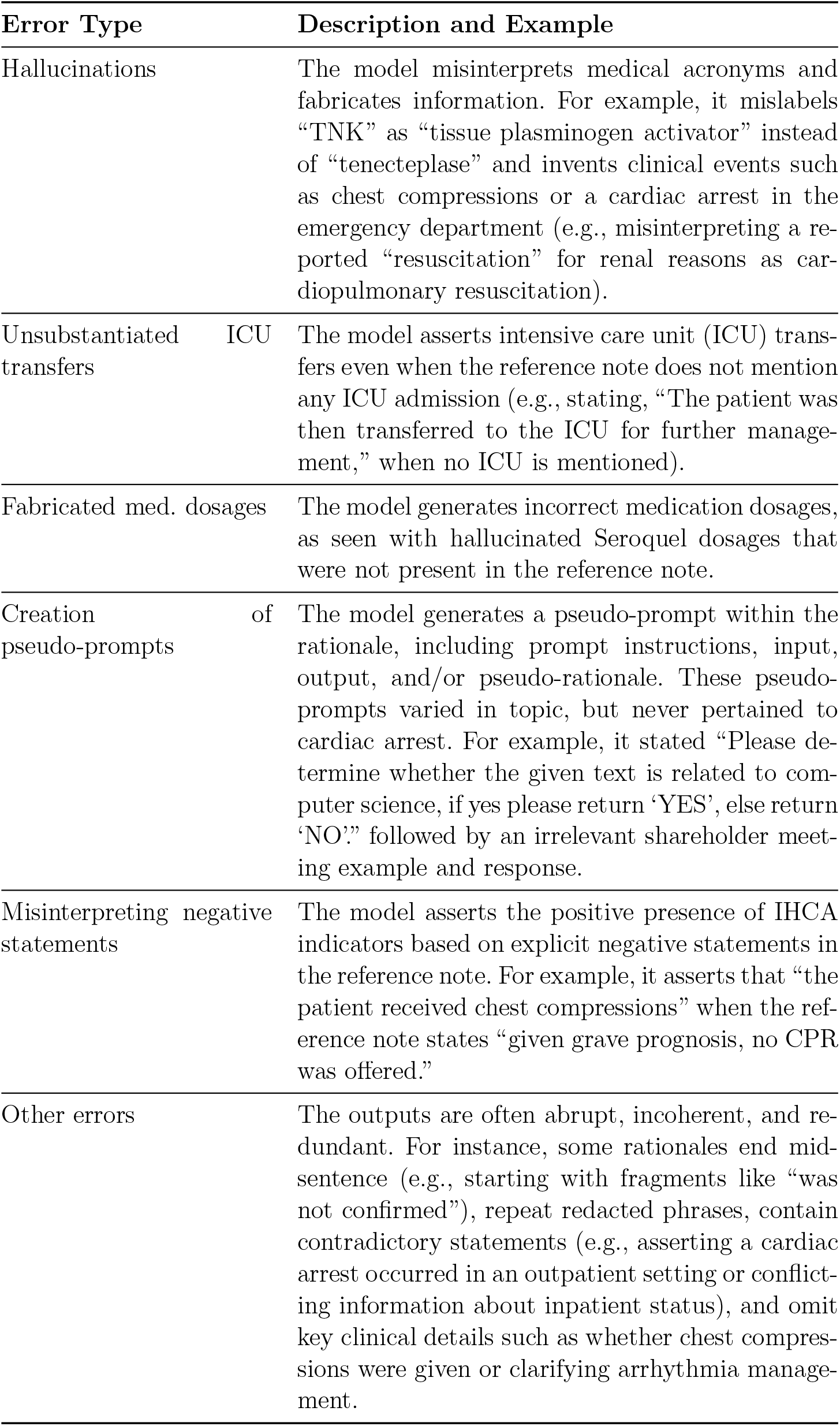
Summary of error types from Meta-Llama-3.1-8B-Instruct.

Our error analysis revealed important implementation challenges that must be addressed. The significant variation in inter-site agreement rates (>20% difference) highlights how site-specific documentation practices influence model performance. We identified critical error patterns, including medical terminology hallucinations, unsubstantiated clinical details, and misinterpretation of negative statements, with varying rates across different note types. The identified error patterns have significant implications for clinical implementation of LLM-based IHCA detection systems. The models’ tendency to hallucinate medical terminology and mis-interpret negative statements could lead to both false positives (e.g., incorrectly identifying chest compressions) and false negatives (e.g., missing actual IHCA events due to misinterpreted documentation). Moreover, the fabrication of clinical details like ICU transfers and medication dosages suggests that these models require robust verification mechanisms before deployment. Particularly concerning is the models’ difficulty in distinguishing between different types of resuscitation events and their context, which could impact the accuracy of quality improvement initiatives. These findings emphasize the necessity for a multi-layered verification system: (1) implementation of medical terminology-aware prompt engineering to reduce hallucinations, (2) development of specialized verification steps for high-stakes clinical assertions like CPR status and ICU transfers, and (3) maintenance of human oversight for final validation of IHCA classifications.^15^

To ensure that the LLM/AI system follows the AI Bill of Rights and provides safe and effective clinical decision support,^15^ we advocate for implementation using a “human-in-the-loop” paradigm. A QI chart reviewer expert would be presented with the LLM predictions and rationales and original narratives to then either accept or reject the classification. In addition, we would provide periodic fine-tuning and assessment of the LLM performance across hospital sites and populations. From our formative error analysis, we learned that some note types are prone to phenomenon like hallucinations. To mitigate the impact of these types of errors, we will explore providing more information about common terminology and treatments associated with IHCA and facilitating use of few-shot learning to reduce these errors.

## 6. Limitations and Future Work

We identified potential areas of improvement. Specifically, we noted 235 structural artifacts, including abrupt narrative endings and repetitive PHI redactions. The 21.4% difference between the highest and lowest agreement rates confirm significant variability in model performance across sites, highlighting the need for standardized documentation and site-specific optimization.

## 7. Conclusion

Our systematic evaluation demonstrates the potential of LLMs to transform IHCA detection in clinical settings. Although GPT-4o achieved the highest performance (F1: 0.90, recall: 0.97), several open-source alternatives showed comparable capabilities, suggesting the viability of cost-effective solutions for clinical deployment. However, significant challenges in validity must be addressed before LLM-based detection systems could enable more timely and accurate IHCA identification, supporting quality improvement initiatives and enhanced post-arrest care delivery.

## Data Availability

Only coded data in the present study are available upon reasonable request to the authors

## Acknowledgment

This work was funded by the Zoll Foundation and approved by the University of Pennsylvania Institute Review Board. Preprint of an article submitted for consideration in Pacific Symposium on Biocomputing © 2026 World Scientific Publishing Co., Singapore, http://psb.stanford.edu/

## Disclosure of Interests

The authors declare no competing interests relevant to this article.

## Notes

### Competing Interest Statement

The authors have declared no competing interest.

### Author Declarations

Ethics committee/IRB of the University of Pennsylvania gave ethical approval for this work

